# Utilization, Determinants and Experiences of Youth-Friendly Sexual and Reproductive Health Services Among Youth in Gurage Zone, Southern Ethiopia: A Concurrent Mixed-Methods Study

**DOI:** 10.64898/2026.08.26.26361385

**Authors:** Getachew Gebreselassie Nida, Sisinyana H. Khunou, David Mphuthi

## Abstract

**Background:** Sexual and reproductive health (SRH) is essential for youth development, particularly in Sub-Saharan Africa where youth represent a significant proportion of the population. Despite global efforts to promote Sexual and Reproductive Health rights, many disadvantaged youths face barriers to accessing comprehensive information and services. Youth-friendly sexual and reproductive health (YFSRH) services are central to improving youth health outcomes. Despite national standards in Ethiopia, implementation challenges persist. Healthcare workers (HCWs) are key actors in promoting and delivering YFSRH services

**Method:** A concurrent mixed-methods design was employed among youth (18–24 years) and health care workers in Guraghe Zone, Southern Ethiopia. Quantitative data were collected using self-administered questionnaires, while qualitative data were gathered through key informant interviews. Quantitative data were analyzed using SPSS version 29, and qualitative data were analyzed thematically.

**Result:** Although youths showed strong interest in Sexual and Reproductive Health information, help-seeking was often delayed due to discomfort, secrecy, social pressure, and limited foresight. Utilization of youth-friendly Sexual and Reproductive Health services was constrained by distance, inconvenient service hours, limited privacy and confidentiality, perceived judgmental provider attitudes, and financial barriers, reducing trust and repeat use.

**Conclusion:** Improving youth Sexual and Reproductive Health requires integrated actions across education, families, and health services. Strengthening multi-channel Sexual and Reproductive Health education with life skills and psychosocial support, alongside decentralized, affordable, confidential, and non-judgmental youth-friendly services, is essential.

## Background

Reproductive health extends beyond the mere absence of disease, encompassing the holistic attainment of optimal physical, mental, and social well-being across all dimensions of the reproductive system functionality (1). SRH is integral to youth development, supported by rights that ensure equality and human dignity (2). Program of action implementation was adopted to protect and promote youth sexual and reproductive health (YSRH) through healthcare, education, information dissemination and efforts to reduce sexually transmitted infections (STIs) and pregnancies (3). SRH is vital for youth development and is protected by sexual and reproductive health rights (SRHR), which emphasis equality and dignity. The youth phase is increasingly recognized as crucial for future health but also as a vulnerable period. Youth aged 15-24, constituting about 20% of the Sub-Saharan African (SSA) population, need to understand the SRH behaviors.

While there have been global efforts to promote SRH, many disadvantaged youth lack autonomies to access comprehensive SRH information and services. The reproductive health needs encompass family planning, HIV/AIDS education, safe sexual behavior, addressing unintended and early pregnancies, sexually transmitted infections (STIs), safe abortion, and safe motherhood. YSRH needs as including family planning, HIV/AIDS education, safe sexual practices, prevention of unintended and early pregnancies, management of STIs, access to safe abortion and support for safe motherhood. Since the ICPD, there has been notable global progress in promoting SRH (4). However, millions of disadvantaged youth lack the autonomy to access comprehensive SRH information and services, due to the taboo attached to the subject (5). Youth face risky behavioral practices such as early sexual initiation, unprotected intercourse and having multiple sexual partners which often lead to adverse SRH outcomes including STIs, HIV/AIDS infections, unwanted pregnancies, and unsafe abortions (6). The purpose of this study is to determine and explore the utilizations of youth-friendly sexual and Reproductive health (YFSRH) services and associated factors among youth in Gurage zone, Southern Ethiopia

## Methods

### Study Setting

The study was conducted in the Gurage Zone, Southern Ethiopia, with Wolkite town as the zonal administrative center, located about 158 km southwest of Addis Ababa. (Fig 1). The zone is administratively divided into 20 districts and 2 town administrations. According to the 2025 projection, it has an estimated population of 1.8 million (7).

**Fig 1.**
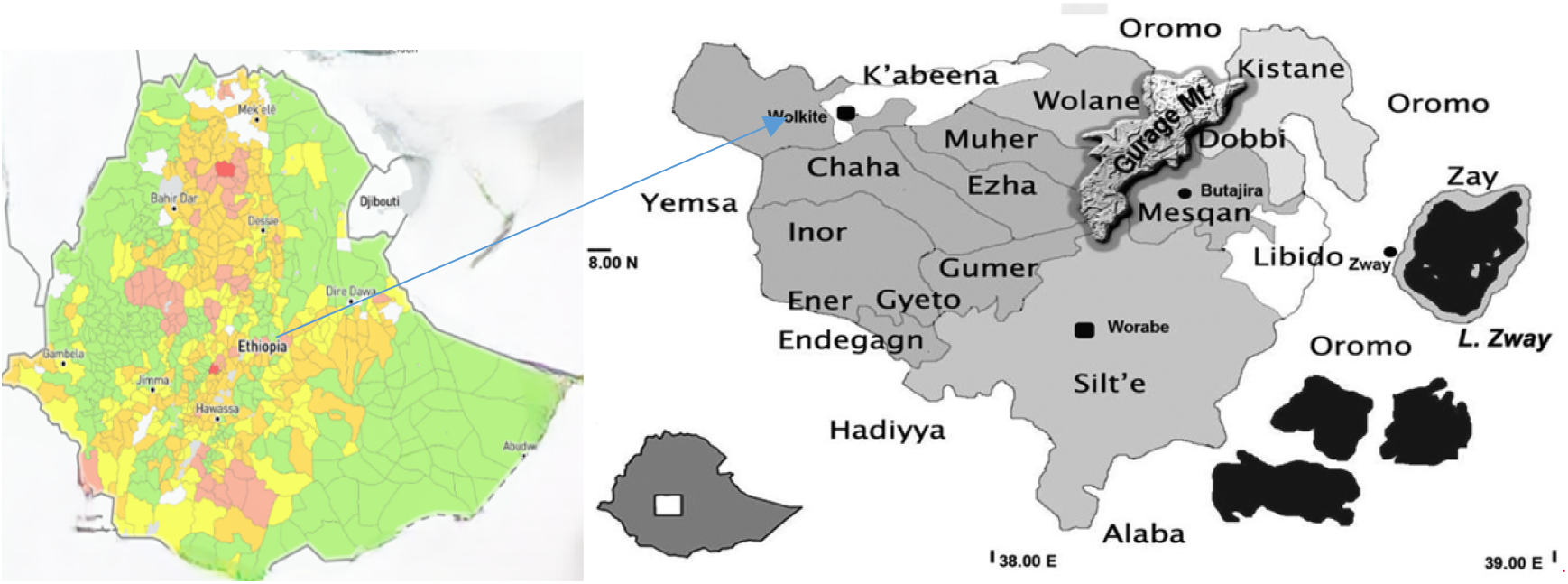
Administrative map of Gurage zone including districts/Woredas. (https://doi.org/10.1371/journal.pone.0198353.g001)

In terms of health infrastructure, Gurage Zone maintains a mixed public and non-governmental health service delivery system. The zone has 7 hospitals, of which five are public including one comprehensive specialized hospital at Wolkite University and one general hospital at Butajira town. Two hospitals are non-governmental or faith-based. In addition, the zone is served by 72 health centers, of which seven are NGO-managed and 402 functional health posts are distributed across the low administrative units.

### Study Design

The mixed methods approach was utilized to collect, examine and incorporate both quantitative and qualitative data to meet the study’s objectives (8). A convergent parallel mixed methods approach was applied.

### Study Population

The study population comprised youth in preparatory school for quantitative phase and health care workers (HCWs) selected from public health facilities under Gurage zone health department to understand the YFSRH services.

### Sample size determination

A two-stage sampling technique was employed to select study participants from the school. In the first stage, students were stratified into two groups based on grade level: Grade 11 and Grade 12. Each grade consisted of four sections labeled A, B, C and D. The number of study participants selected from each grade was proportionally allocated according to the size of the respective sections.

In the second stage, students within each section were selected using systematic random sampling. The list of students in each section served as the sampling frame. After determining the sampling interval, the first participant was chosen using a lottery method. Subsequent participants were then selected by adding the calculated sampling interval until the required sample size for each section was achieved.

For the qualitative phase, purposive sampling was used to select Youth-Friendly Sexual and Reproductive Health (YFSRH) service centers. This approach relies on the researcher’s knowledge of the population to deliberately select participants or sites that are most relevant to the research objectives (9).

## Sampling procedure

The sample size for the quantitative component was determined using the single population proportion formula. A proportion of 50% was assumed for youth utilization of Youth-Friendly Sexual and Reproductive Health (YFSRH) services [8], with a 5% margin of error and a 95% confidence interval (Zα/2 = 1.96). After adding a 10% allowance for non-response, the initial sample size of 384 was increased to a final total of 422 participants. The study was conducted as a school-based descriptive survey among youth aged 18–24 years.

For the qualitative phase, twelve YFSRH service centers were purposively selected from the fifteen public health facilities available in the study area.

## Data collection method and tools

Quantitative structured self-administered questionnaires under each section were arranged in a way that maintained a flow of the discussion. Response options were checked for exhaustiveness. These options were arranged to maintain flow and order in the ranking. Survey questions were largely based on structured yes or no response options and Likert scale options. For some of the questions, a list of possible response options was provided, and an additional response option called “other” was provided for those response options out of the listed.

The timing and setting for administering the questionnaire were coordinated with the school principal to ensure youth in school could focus and feel comfortable. Preparations included organizing printed questionnaires and writing tools. Youth in school were briefed on the purpose of the questionnaire, the importance of honest responses, and confidentiality. Instructions on completing the questionnaire were provided, and any questions were addressed. The questionnaires were distributed, and school youth were given one and half hours to complete them. Completed questionnaires were promptly collected and securely stored until processing.

For the qualitative phase data collection process, the list of active health facilities that has been providing YFSR and the respective health facilities focal persons, were obtained from the zonal health department Potential participants were contacted via email and phone, where they received a brief overview of the study and an invitation to participate ahead of the face to face interview. Arrangements were made with participants to schedule interviews at convenient times, and all necessary materials, including audio recording devices and consent forms, were prepared. A quiet, private location was chosen to ensure confidentiality and comfort. The interviews were conducted by the researcher using a semi-structured interview guide.

## Data quality assurance

The tools were initially prepared in English, translated in to local language (Amharic) and then translated back to English to check for consistency. Before data collection, the tools were pre-tested among school base youths and health care workers (HCWs) who were not included in the study, 10% of the sample were taken for the pre-test. The pre-test analysis revealed minor issues that need addressing including errors in question order, spelling mistakes, missing information and the time needed to complete the self-administered questionnaire. The tool was amended in alignment with identified gaps and limitations during pre-testing.

## Data entry and analysis

Quantitative data were coded and analyzed using SPSS version 28 to generate descriptive statistics (frequency, percentage, mean, median, standard deviation) and to examine relationships using binary logistic regression. Variables with p < 0.25 in univariable analysis were included in multivariable analysis, and those with p < 0.05 at 95% CI were considered determinants.

Qualitative data were transcribed, organized and manually analyzed using thematic analysis, with audio recordings securely stored and labeled to ensure systematic interpretation and contextual understanding.

## Ethical consideration

Ethical clearance was obtained from the departmental higher degree research and ethics committee of University of South Africa (UNISA). A letter of permission from Guraghe zone health department was granted to undertake the research in the selected preparatory schools and health facilities providing youth sexual and reproductive health in the study area. The respondents were informed about the objective and purpose of the study. Oral and written consent was obtained from each respondent before administering the questionnaire. To assure confidentiality no name or personal identifying information was written on the questionnaire and information was recorded anonymously

## Results

### Socio-demographic information for the quantitative phase

The sociodemographic data (**Table 1)** show that 50.2% males (n=212) and 49.8% females (n=210) participated in the study, indicating nearly equal gender representation. The majority of participants were youth, with 84.8% (n=358) aged between 18-20 years, and 15.2% (n=64) aged 21-23 years, suggesting a focus on youth, likely from secondary education.

**Table 1.**
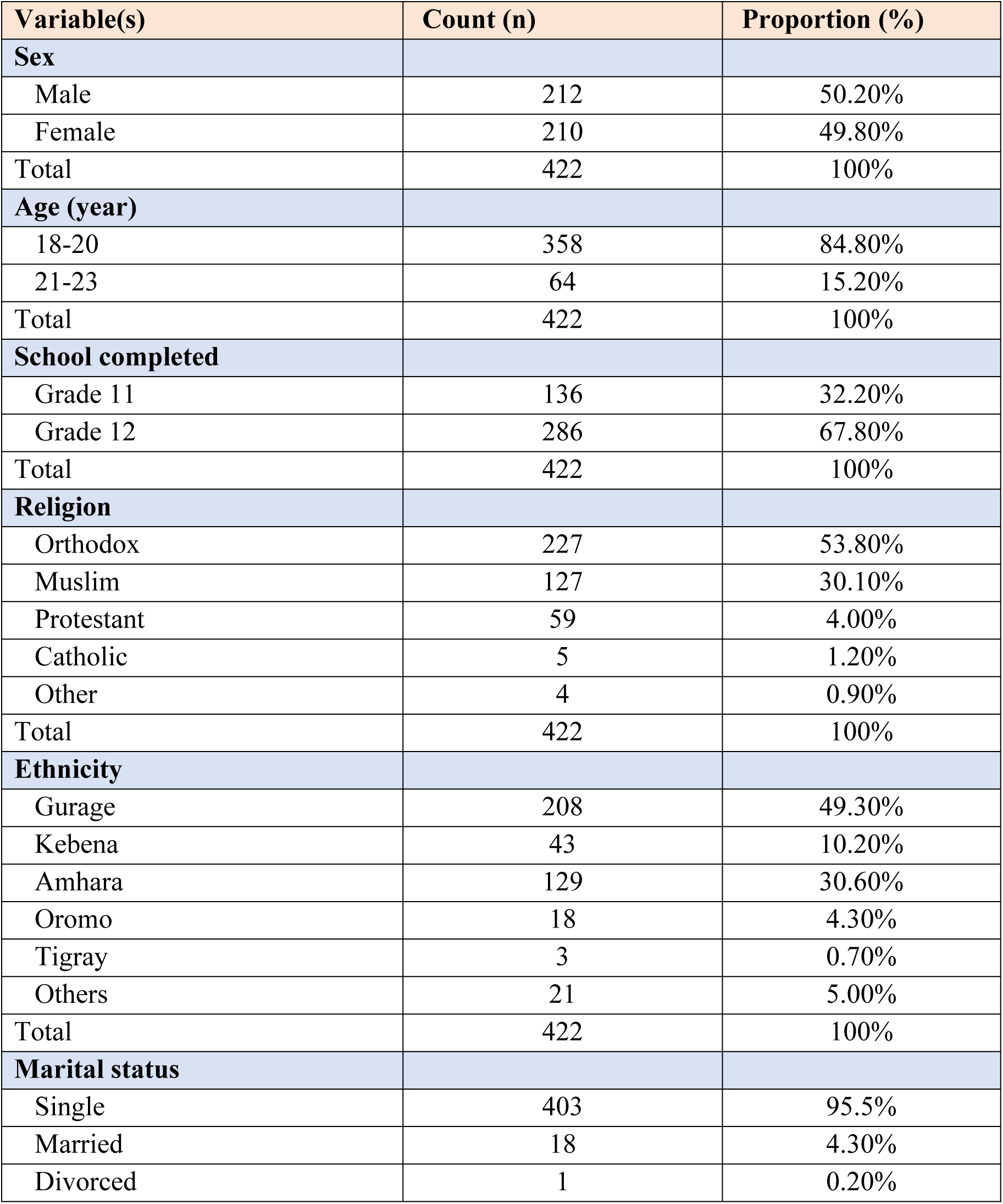
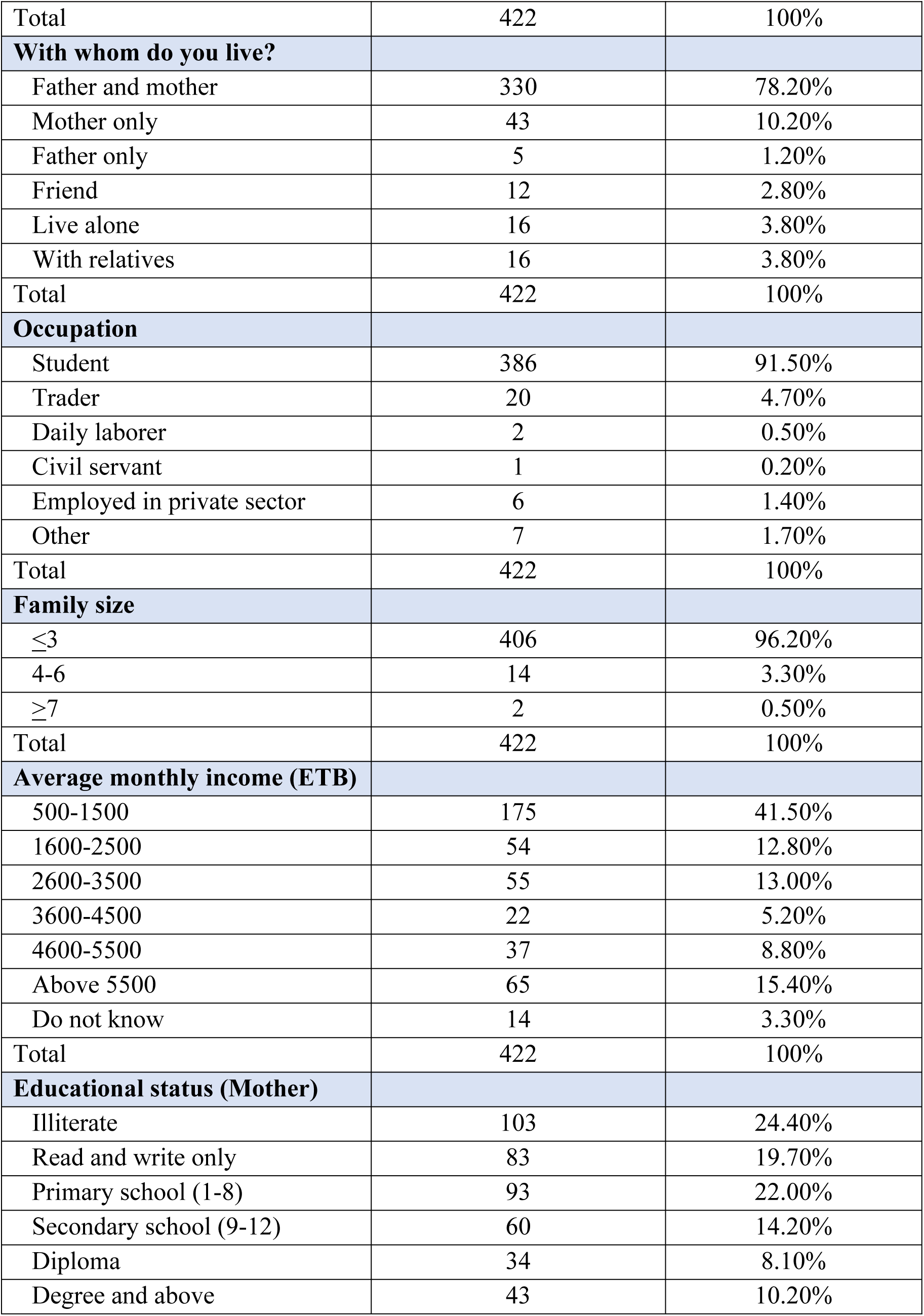
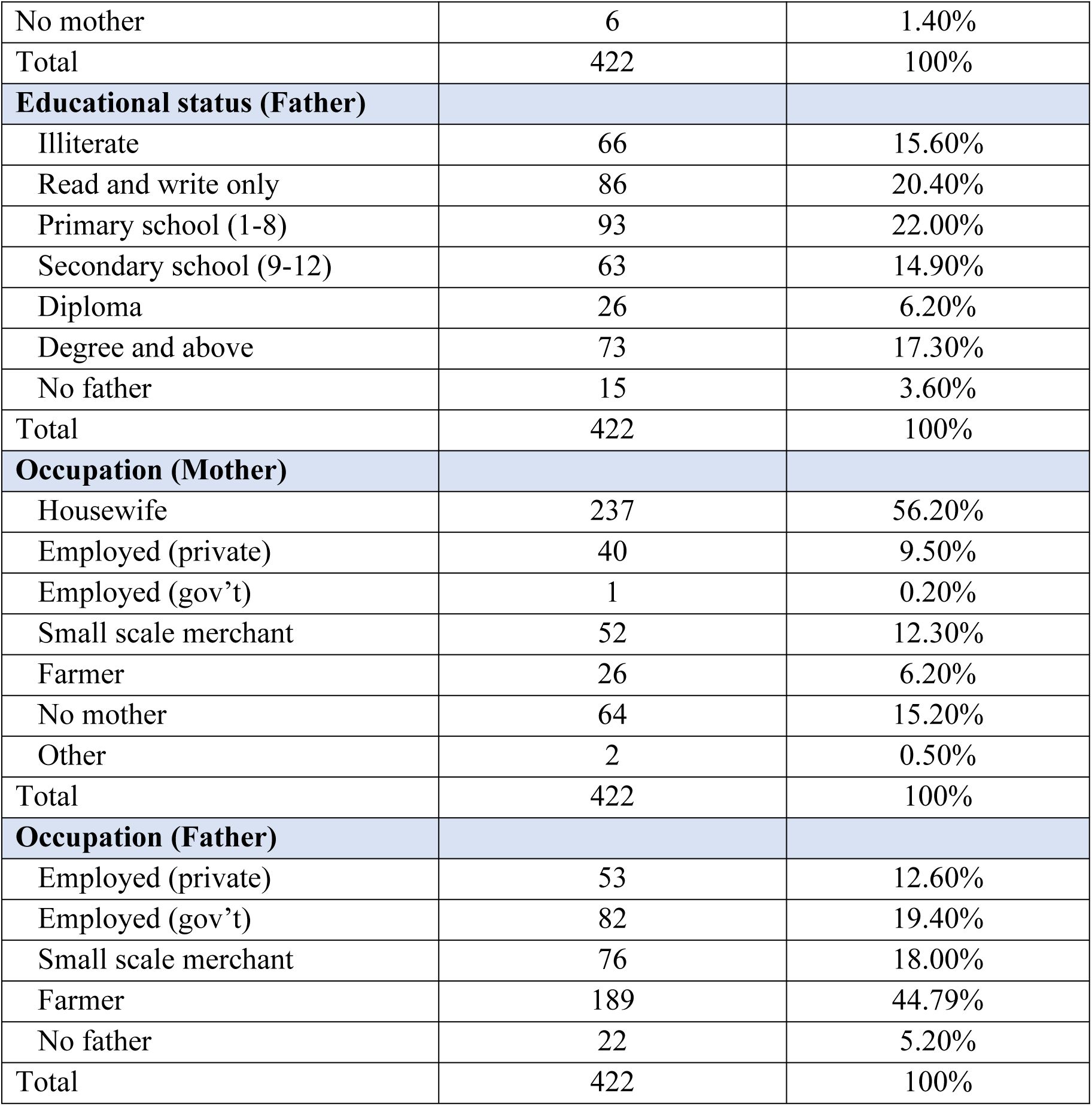
Socio-demographic characteristics of the participants in Gurage zone, Central Ethiopia Region (n =422)

| Variable(s) | Count (n) | Proportion (%) |
| --- | --- | --- |
| **Sex** |  |  |
| Male | 212 | 50.20% |
| Female | 210 | 49.80% |
| Total | 422 | 100% |
| **Age (year)** |  |  |
| 18-20 | 358 | 84.80% |
| 21-23 | 64 | 15.20% |
| Total | 422 | 100% |
| **School completed** |  |  |
| Grade 11 | 136 | 32.20% |
| Grade 12 | 286 | 67.80% |
| Total | 422 | 100% |
| **Religion** |  |  |
| Orthodox | 227 | 53.80% |
| Muslim | 127 | 30.10% |
| Protestant | 59 | 4.00% |
| Catholic | 5 | 1.20% |
| Other | 4 | 0.90% |
| Total | 422 | 100% |
| **Ethnicity** |  |  |
| Gurage | 208 | 49.30% |
| Kebena | 43 | 10.20% |
| Amhara | 129 | 30.60% |
| Oromo | 18 | 4.30% |
| Tigray | 3 | 0.70% |
| Others | 21 | 5.00% |
| Total | 422 | 100% |
| **Marital status** |  |  |
| Single | 403 | 95.5% |
| Married | 18 | 4.30% |
| Divorced | 1 | 0.20% |
| Total | 422 | 100% |
| **With whom do you live?** |  |  |
| Father and mother | 330 | 78.20% |
| Mother only | 43 | 10.20% |
| Father only | 5 | 1.20% |
| Friend | 12 | 2.80% |
| Live alone | 16 | 3.80% |
| With relatives | 16 | 3.80% |
| Total | 422 | 100% |
| **Occupation** |  |  |
| Student | 386 | 91.50% |
| Trader | 20 | 4.70% |
| Daily laborer | 2 | 0.50% |
| Civil servant | 1 | 0.20% |
| Employed in private sector | 6 | 1.40% |
| Other | 7 | 1.70% |
| Total | 422 | 100% |
| **Family size** |  |  |
| $\leq 3$ | 406 | 96.20% |
| 4-6 | 14 | 3.30% |
| $\geq 7$ | 2 | 0.50% |
| Total | 422 | 100% |
| **Average monthly income (ETB)** |  |  |
| 500-1500 | 175 | 41.50% |
| 1600-2500 | 54 | 12.80% |
| 2600-3500 | 55 | 13.00% |
| 3600-4500 | 22 | 5.20% |
| 4600-5500 | 37 | 8.80% |
| Above 5500 | 65 | 15.40% |
| Do not know | 14 | 3.30% |
| Total | 422 | 100% |
| **Educational status (Mother)** |  |  |
| Illiterate | 103 | 24.40% |
| Read and write only | 83 | 19.70% |
| Primary school (1-8) | 93 | 22.00% |
| Secondary school (9-12) | 60 | 14.20% |
| Diploma | 34 | 8.10% |
| Degree and above | 43 | 10.20% |
| No mother | 6 | 1.40% |
| Total | 422 | 100% |
| **Educational status (Father)** |  |  |
| Illiterate | 66 | 15.60% |
| Read and write only | 86 | 20.40% |
| Primary school (1-8) | 93 | 22.00% |
| Secondary school (9-12) | 63 | 14.90% |
| Diploma | 26 | 6.20% |
| Degree and above | 73 | 17.30% |
| No father | 15 | 3.60% |
| Total | 422 | 100% |
| **Occupation (Mother)** |  |  |
| Housewife | 237 | 56.20% |
| Employed (private) | 40 | 9.50% |
| Employed (gov't) | 1 | 0.20% |
| Small scale merchant | 52 | 12.30% |
| Farmer | 26 | 6.20% |
| No mother | 64 | 15.20% |
| Other | 2 | 0.50% |
| Total | 422 | 100% |
| **Occupation (Father)** |  |  |
| Employed (private) | 53 | 12.60% |
| Employed (gov't) | 82 | 19.40% |
| Small scale merchant | 76 | 18.00% |
| Farmer | 189 | 44.79% |
| No father | 22 | 5.20% |
| Total | 422 | 100% |

Regarding educational background, 67.8% (n=286) were Grade 12, and 32.2% (n=136) were Grade 11. The religious affiliation was diverse, with 53.8% (n=227) identifying as Orthodox Christians, 30.1% (n=127) as Muslims, 4.0%(n=59) as Protestants, and smaller groups including Catholics 1.2% (n=5) and Adventists 0.9%(n=4).

Ethnicity, 49.3% (n=208) of participants were Gurage, followed by 30.6% (n=129) Amhara, and 10.2% (n=43) Kebena, with other smaller ethnic groups included. Regarding the marital status, the majority (95.5%) were single, with only 4.3%(n=18). married and 0.2%(n=1). divorced, reflecting the youth of the sample.

Regarding living arrangements, 78.2% (n=330) lived with both parents, while smaller proportions lived with only one parent 10.2%(n=43) with mother, 1.2%(n=5) with father), friends 2.8% (n=12), relatives 3.8% (n=16). or alone 3.8% (n=16). Finally, a high number of participants, 91.5% (n=386) of participants were students, with small numbers involved in trade 4.7% (n=20), daily labor 0.5% (n=2), civil service 0.2% (n=1) and private sector work 1.4% (n=6).

### Sexual and Reproductive Health Knowledge and Behaviors

#### Sources of SRH Information

Participants reported multiple sources of SRH information, with schools and teachers being the most frequently cited (28.0%), followed by media (26.8%) and health workers (17.5%). Peers and family members were less commonly reported **(Table 2).**

**Table 2.** Sexual and reproductive health knowledge and perception of youth (n =422)

| Variable(s) | Sample (n) | Proportion (%) |
| --- | --- | --- |
| Source of information about SRH matter |  |  |
| School/Teachers | 118 | 28.00% |
| Media | 113 | 26.80% |
| Home | 47 | 11.10% |
| Peers | 57 | 13.50% |
| Health workers | 74 | 17.50% |
| Other | 13 | 3.10% |
| Total | 422 | 100% |
| Have you ever had sexual intercourse? |  |  |
| Yes | 90 | 21.30% |
| No | 332 | 78.70% |
| Total | 422 | 100% |
| First age (year) of sexual intercourse |  |  |
| 18-20 | 84 | 19.90% |
| 21-23 | 2 | 0.50% |
| No response | 336 | 79.60% |
| Total | 422 | 100% |
| Number of partners in last 12 months |  |  |
| Never | 369 | 87.40% |
| Only one partner | 41 | 9.70% |
| Two to three partners | 7 | 1.70% |
| > 3 partners | 5 | 1.20% |
| Total | 422 | 100% |
| With whom did you have sex? |  |  |
| Friend | 35 | 8.30% |
| Teacher | 26 | 6.20% |
| Commercial sex worker | 2 | 0.50% |
| Fiancé | 9 | 2.10% |
| Girlfriend | 7 | 1.70% |
| Relative | 8 | 1.90% |
| Forced intercourse/rape | 1 | 0.20% |
| Other | 8 | 1.90% |
| Don't remember | 34 | 8.10% |
| Reason for commencing sexual intercourse |  |  |
| Having sexual desire | 35 | 8.30% |
| Peer pressure | 26 | 6.20% |
| Teacher pressure | 2 | 0.50% |
| Friends all doing it | 9 | 2.10% |
| Felt I was the right age | 7 | 1.70% |
| Substance use | 8 | 1.90% |
| Economic benefit | 1 | 0.20% |
| Raped | 8 | 1.90% |
| Other | 34 | 8.10% |
| Duration of partnership before 1<sup>st</sup> sexual |  |  |
| Met for a few weeks | 7 | 1.70% |
| For a few months | 14 | 3.30% |
| For a few years | 25 | 5.90% |
| In a committed relationship | 18 | 4.30% |
| Living together as a couple | 3 | 0.70% |
| Engaged to be married | 4 | 0.90% |
| Married | 10 | 2.40% |
| Other | 31 | 7.30% |
| Use of contraceptive during 1<sup>st</sup> sexual intercourse |  |  |
| Yes | 39 | 9.20% |
| No | 54 | 12.80% |
| Don't remember | 28 | 6.60% |
| Type of contraceptive used |  |  |
| Withdrawal (pulling out) | 7 | 1.70% |
| Condom only | 25 | 5.90% |
| Birth control pills | 12 | 2.80% |
| Emergency pills | 8 | 1.90% |
| Injectable | 9 | 2.10% |
| IUD | 7 | 1.70% |
| Natural family planning | 35 | 8.30% |
| Other | 2 | 0.50% |
| Frequency of condom use |  |  |
| Occasionally | 32 | 7.60% |
| Always | 29 | 6.90% |
| Consistently | 11 | 2.60% |
| Reason for not using condom |  |  |
| Male partner objection | 8 | 1.90% |
| Embarrassed to buy | 16 | 3.80% |
| I trust my partner | 25 | 5.90% |
| I was drunk | 12 | 2.80% |
| It reduces my sexual pleasure | 15 | 3.60% |
| Other | 26 | 6.20% |
| Availability of boyfriend/girl friend |  |  |
| Yes | 281 | 66.60% |
| No | 141 | 33.40% |
| Total | 422 | 100% |

Qualitative findings reinforced the central role of schools as structured platforms for delivering SRH information:

> “*Schools are a major focus, and topics like YFSRH are prioritized in health education curricula… we conduct youth-friendly services in schools and provide counselling” (HCW; P7).*

Peers were identified as influential but inconsistent sources of information:

> *“Some peers promote awareness… but others spread negativity… this stigma affects her” (HCW; P6).*

### Sexual Practices and Behavioral Patterns

As shown Table 2, approximately 21.3% of respondents reported having engaged in sexual intercourse, with most initiating between the ages of 18 and 20. Among sexually active youth, the majority reported either no partner or a single partner in the past 12 months.

Contraceptive use at first sexual intercourse was low (9.2%), and condom use was inconsistent. Reported reasons for non-use included partner objection, embarrassment, trust in partners, and perceived reduction in sexual pleasure.

Qualitative findings provided insight into the social drivers of these behaviors, particularly peer influence:

> *“My friend did it, so why shouldn’t I? This mindset is driven by peer pressure” (HCW; P5).*

### Awareness of YFSRHS

Most respondents (68.5%) reported having information about YFSRHS, while nearly one-third (31.5%) lacked awareness. Teachers and peers were the primary sources of information (Table 2).

Qualitative findings suggest that awareness is largely driven by school-based outreach and community engagement, although gaps persist among rural and out-of-school youth.

### Perceived Importance of YFSRHS

A large majority of respondents (81.7%) perceived YFSRHS as important for improving youth health. Qualitative findings support this, indicating that youth value services that are accessible, respectful, and confidential.

### Youth-Friendly Sexual and Reproductive Health (YFSRH) Services

Youth-Friendly Sexual and Reproductive Health (YFSRH) services aim to provide confidential, accessible, respectful, and developmentally appropriate care, recognizing youth as a critical stage for establishing lifelong health behaviors.

However, despite awareness of available services, many youths hesitate to utilize them due to fear of judgment, lack of privacy, and provider insensitivity, highlighting the need for holistic, youth-centered approaches that strengthen confidentiality, provider competence, and youth participation in program design and evaluation.

### Accessibility of YFSRH services

The quantitative results verify that only 20.4% of respondents found the YFSRH facility easily accessible, while 79.6% reported difficulty reaching services. More than half (56.5%) indicated that the nearest facility was at least an hour’s walk from their residence, and 87% stated that the facilities were not open during times convenient for them. Accessibility was further constrained by cost, with 84.4% reporting that YFSRH services were not free or affordable. Overall, the quantitative data indicate that most youth face multiple barriers to accessing SRH services, including geographic distance, inconvenient service hours, and financial limitations **(Table 3).**

**Table 3.** Youth-friendly sexual and reproductive health (YFSRH) and Associated factors (n =422)

| Variable(s) | Count (n) | Proportion (%) |
| --- | --- | --- |
| Respondents who have information about YFRHS |  |  |
| Have information | 289 | 68.50% |
| Don't have information | 133 | 31.50% |
| Total | 422 | 100% |
| Source of information about YFSRHS |  |  |
| Parents | 82 | 20.40% |
| Peers | 108 | 26.90% |
| Teachers | 138 | 34.40% |
| Health Workers | 94 | 22.27% |
| Total | 422 | 100% |
| Distance to the facility |  |  |
| Near, short walking distance | 85 | 21.60% |
| 30 minutes round walking distance | 86 | 21.90% |
| Far, one hour and above round walking distance | 222 | 56.50% |
| Essential SRH services known and utilized in the facility |  |  |
| Contraception/condom | 102 | 24.40% |
| VCT | 98 | 23.40% |
| STIs prevention and management | 55 | 13.20% |
| Treatment of STIs | 38 | 9.10% |
| Care of young pregnant and ANC | 37 | 8.9% |
| Pregnancy test | 28 | 6.70% |
| Counselling service on safe sex | 15 | 3.60% |
| Prevention and management of gender-based violence | 14 | 3.30% |
| Prevention of unsafe abortion and management of post-abortion care | 5 | 1.20% |
| Don't know | 30 | 7.11% |
| Total | 422 | 100% |
| Perception on YFRH services in improving youths' health |  |  |
| Yes | 344 | 81.70% |
| No | 78 | 18.48% |
| Total | 422 | 100% |
| Factors that influenced the utilizations of YFRHS services |  |  |
| Mass-media messages | 275 | 68.90% |
| Advice from others | 94 | 22.27% |
| Illness of close relative due to HIV or STI | 29 | 7.30% |
| Death of close relative due to HIV, STI or abortion | 24 | 6.00% |
| Total | 422 | 100% |
| Reasons for not using YFRHS |  |  |
| Religious matters | 189 | 46.70% |
| Cultural matters | 153 | 37.80% |
| I don't know | 80 | 18.96% |
| Total | 422 | 100% |
| Respondents who visited health facility and missed the services |  |  |
| Yes | 108 | 25.70% |
| No | 314 | 74.41% |
| Total | 422 | 100% |
| Reasons for missing YFFRH services |  |  |
| Lack of privacy at the facility | 51 | 12.10% |
| Lack of money for the service | 43 | 10.20% |
| I found neighbors and felt ashamed | 60 | 14.20% |
| Service providers were harsh and denied service | 171 | 40.50% |
| The clinic was closed | 63 | 14.90% |
| The time of service provision is inconvenient | 15 | 3.60% |
| The queue was long | 19 | 4.50% |
| Total | 422 | 100% |
| Accessibility of the YFSRHS facility |  |  |
| Yes | 86 | 20.40% |
| No | 336 | 79.60% |
| Total | 422 | 100% |
| Facility open during hours that are convenient for youth |  |  |
| Yes | 55 | 13.00% |
| No | 367 | 87.00% |
| Total | 422 | 100% |
| Specific clinic times or spaces set aside for youth |  |  |
| Yes | 59 | 14.00% |
| No | 363 | 86.00% |
| Total | 422 | 100% |
| YSRHS offered for free or at rates affordable |  |  |
| Yes | 66 | 15.60% |
| No | 356 | 84.40% |
| Total | 422 | 100% |
| Are there short waiting times for youth to get an appointment before talking to a health provider? |  |  |
| Yes | 63 | 15.17% |
| No | 358 | 84.83% |
| Total | 422 | 100% |
| There is a separate discreet entrance for youth to ensure their privacy |  |  |
| Yes | 59 | 13.98% |
| No | 363 | 86.02% |
| Total | 422 | 100% |
| Counselling and treatment rooms allow for privacy (both visual and auditory) |  |  |
| Yes | 75 | 17.77% |
| No | 347 | 82.23% |
| Total | 422 | 100% |
| Transparent and confidential mechanism for youth to submit complaints or feedback about SRHS at the facility? |  |  |
| Yes | 68 | 16.11% |
| No | 354 | 83.89% |
| Total | 422 | 100% |
| SRH educational materials and posters available on site |  |  |
| Yes | 54 | 12.80% |
| No | 368 | 87.20% |
| Total | 422 | 100% |
| Reasons for not using youth friendly services in the facilities |  |  |
| Didn't start the sexual life | 152 | 36.02% |
| The public opinion disagrees | 67 | 15.88% |
| The attitude and behavior medical staff | 77 | 18.25% |
| Only married people need YFS | 68 | 16.11% |
| Parents, parent's friends and teachers could see the youth | 27 | 6.87% |
| Shame, fear of gynecological examination | 19 | 4.50% |
| Contraceptives' price | 1 | 0.24% |
| Using natural contraception | 5 | 1.18% |
| The lack of information | 4 | 0.95% |
| Getting SRHS without permission from my parent or guardian |  |  |
| Total | 422 | 100% |
| Agree | 193 | 50.70% |
| Disagree | 188 | 49.30% |
| Confidentiality is respected at YFRH facility |  |  |
| Agree | 159 | 44.80% |
| Disagree | 196 | 55.20% |
| Feel comfortable at YFRH facility |  |  |
| Agree | 140 | 39.20% |
| Disagree | 217 | 60.80% |
| Clinic staff are respectful and friendly |  |  |
| Agree | 124 | 34.30% |
| Disagree | 238 | 65.70% |
| Health provider at the clinic are non-judgmental |  |  |
| Agree | 115 | 32.20% |
| Disagree | 242 | 67.80% |
| Culture and identity were understood and valued |  |  |
| Agree | 132 | 36.30% |
| Disagree | 232 | 63.70% |
| Staff respect customers and their concerns |  |  |
| Agree | 140 | 37.90% |
| Disagree | 229 | 62.10% |
| Everything was explained before it happened |  |  |
| Agree | 147 | 39.70% |
| Disagree | 223 | 60.30% |
| Felt comfortable asking questions |  |  |
| Agree | 149 | 40.10% |
| Disagree | 223 | 59.90% |
| Total | 422 | 100% |

**Table 4.** Themes, categories and sub-categories that emerged from HCWs interview.

| Theme | Sub-theme | Category (Code) |
| --- | --- | --- |
| 1. YFSRH Service Accessibility and Design | 1.1. Proximity to YFSRH facility | 1.1.1. Accessibility for YFSRH |
|  |  | 1.1.2. Dedicated youth-friendly space |
|  |  | 1.1.3. Flexible Service hours for youth |
|  |  | 1.1.4. School-based programme |
|  | 1.2. The support required from the HCW | 1.2.1. Friendly provider approach |
|  |  | 1.2.2. Confidentiality and privacy assurance |
|  | 1.3. Supportive structure and stakeholders | 1.3.1. Support from health extension workers |
|  |  | 1.3.2. School clubs and teacher collaboration |
|  |  | 1.3.3. Engagement by trained focal persons |
|  |  | 1.3.4. Peer and partner referrals |

The qualitative findings confirmed and enriched these quantitative patterns. HCWs emphasized that long distances, lack of dedicated youth spaces, and rigid operating hours limit youth access to SRH services. Several participants highlighted that youth living in rural areas must walk long distances or rely on costly transportation to reach a health center. HCW explained **(Table 4):**

> *“Access is a significant issue; many youths live in rural areas and must travel uncomfortable distances, sometimes up to two hours on foot, to reach our facility” **(HCW; P4).***

Similarly, other participants indicated that accessibility is a challenge for the youth to utilise the YFSRH:

> *“Those from distant villages don’t come to us. They stay there until complications arise, then come for abortions. They hide pregnancies, and when problems occur, they come. If education were integrated into every health cell or if there were a dedicated centre, as I said earlier, access would improve. But I wouldn’t say it’s fully accessible” **(HCW; P6)***

There is a strong convergence between the quantitative and qualitative findings. Both confirm that youth face significant physical, financial, and organizational barriers in accessing YFSRH services. Quantitative data quantify the extent of inaccessibility, while qualitative narratives provide insight into the lived challenges and potential remedies.

### Confidentiality and privacy

The quantitative findings indicate that only 44.8% of youth agreed that confidentiality was respected when receiving SRH services, while 55.2% disagreed. In addition, 83% reported a lack of privacy in counselling or treatment rooms **(Table 3).** These results suggest that issues of confidentiality and privacy remain critical barriers to trust and service utilizations among youth. Qualitative data reinforced these results by demonstrating how provider attitudes, inadequate infrastructure, and cultural norms undermine privacy. Health workers stressed that maintaining confidentiality is central to youth trust and service continuity. HCW stated **(Table 4).**

> *“The provider must assure them that confidentiality will be maintained between the youth and the professional, with no fear involved. Youth should feel fully accepted, and the provider should deliver services transparently” **(HCW; P5).***

In agreement with other participants:

> *“There’s no breach here. Some youth feel their privacy is at risk because the community knows they visited, but it’s just their perception. For instance, if a neighbor sees them enter, they might assume it’s for family planning. But medically, there’s no exposure. If a client says, ‘Keep this between us,’ it’s their right. We tell them, only you and I will know” **(HCW; P9).***

However, some HCWs acknowledged that physical layouts of health facilities often expose youth to observation by other clients, discouraging them from seeking care. Others mentioned that in small communities, fear of gossip or judgment from staff further inhibits youth from accessing SRH services. Both results converge strongly in recognizing confidentiality and privacy as critical determinants of youth trust in SRH services.

### Provider attitude and non-discrimination

The survey showed that 65.7% of youth perceived clinic staff as unfriendly, 67.8% reported judgmental behavior, and 63.7% felt that their cultural identity was not respected during SRH service interactions **(Table 3).** These results reflect persistent attitudinal barriers that hinder youth from seeking services. The qualitative findings provided a more nuanced view of provider attitudes. HCWs recognized that non-judgmental engagement, empathy, and friendliness are essential to encourage youth service utilization. The qualitative data also highlighted that cultural and generational gaps between providers and youth can contribute to misunderstandings and negative perceptions. One participant remarked **(Table 4)**

> *“They need to be approached in a friendly way.” (HCW; P10).*

Others acknowledged that judgmental attitudes by some colleagues discourage youth:

> *“Some professionals might restrict her freedom to access care, thinking, why is this girl here? What is she trying to do?” (HCW; P5)*

The integration affirms strong convergence between quantitative and qualitative findings. The numerical findings quantify youth dissatisfaction with provider behavior while qualitative narratives expose the underlying reasons, moral judgment, lack of empathy, and cultural insensitivity. This convergence underscores that provider attitude is a decisive determinant of youth SRH service uptake.

### Affordability of YFSRH services

The quantitative results showed that 84.4% of respondents reported that youth-friendly YFSRH services were unaffordable or not free **(Table 3).** This financial barrier was one of the most frequently mentioned deterrents to accessing care. Many youths indicated that they could not afford transportation costs, consultation fees, or service-related expenses such as laboratory tests and contraceptive commodities. Qualitative findings reinforced this observation. HCWs noted that financial hardship was a major deterrent preventing youth from seeking care. Participants explained that the costs associated with traveling to health facilities and paying for services discouraged many youths, especially those still in school or unemployed. HCW stated **(Table 4):**

> *“Getting the youth friendly centers incurs costs, many cannot afford.” (HCW; P5).*

Another added that even when services were officially free, hidden costs such as transport, waiting time or required materials still excluded the poorest youth from access. The two data sets converge strongly, both emphasizing that financial constraints represent a systemic barrier to equitable SRH access. Quantitative results measure the magnitude of the problem, while qualitative narratives clarify that affordability issues extend beyond direct service fees to include transportation costs, opportunity costs, and economic dependency on parents. The triangulation underscores the importance of policy-level interventions to ensure that youth services are truly free and financially accessible. Health systems must priorities subsidized or community-based SRH services, school-based outreach, and voucher schemes to reduce cost-related barriers.

### Youth-centeredness and feedback mechanisms

In the quantitative data, 84.5% of respondents reported that no feedback or complaint mechanism existed in the facilities they attended, and 63.7% disagreed that their cultural identity was valued. This indicates a lack of structured channels through which young people can express their concerns, contribute to service design, or provide feedback about their experiences. The qualitative data echoed these findings. Many noted the absence of youth-specific spaces and limited avenues for youth to participate in service planning or evaluation. They also acknowledged that youth services often remain embedded within general health service frameworks, with little differentiation or youth input. This is explained in the vignette below **(Table 4):**

> *“If providers offered all services in one place, like a welcoming center, access would improve.” (HCW; P7).*

HCWs also emphasized that creating a youth corner within facilities and establishing regular forums for youth feedback would enhance satisfaction and encourage return visits. Both results converge on the need for greater youth participation and voice in SRH services. Quantitative findings indicate the absence of institutional mechanisms for feedback, while qualitative insights highlight how this exclusion limits service responsiveness and trust.

### Comprehensive information and education

The quantitative results show that 68.5% of youth reported having some information about youth-friendly SRH services, while 31.5% lacked awareness. Moreover, 87.2% stated that no educational materials were available at the facilities. The main sources of information were teachers (34.4%) and peers (26.9%) **(Table 3).** These findings suggest that while some awareness exists, the quality and consistency of SRH information are limited. Qualitative data further emphasizes the role of education in shaping youth awareness. Health workers described school-based program and peer-led sessions as effective in promoting SRH knowledge. Educational efforts are essential in rural areas where media access is limited. HCW mentioned **(Table 4):**

> *“Schools are a major focus… we provide counselling.” (HCW; P7).*

Another explained that educating youth about SRH early helps prevent risky behavior and dispel myths surrounding sexuality and reproduction. There is a clear convergence between the quantitative and qualitative findings. Both indicate that schools and peers serve as the most common sources of SRH information, whereas health facilities are underutilized for education. Combined finding reveals that the absence of consistent IEC materials at health facilities contributes to gaps in SRH knowledge

### Utilisation factors and barriers

The quantitative results showed that 68.9% of respondents cited mass media as a major source influencing their SRH awareness. However, 46.7% reported religion and 37.8% cited cultural norms as barriers to using YFSRH services. Additionally, 40.5% reported experiencing harsh treatment from providers, further reducing service utilizations **(Table 3)**. Qualitative findings supported these results, revealing that religious and cultural beliefs, negative provider behavior and limited outreach were the most significant barriers to service uptake. One HCW stated:

> *“Those from distant villages don’t come until complications arise.” (HCW; P6).*

HCWs also described how conservative community values discourage open discussions about sexuality and contribute to youth hesitation in seeking care. Integration demonstrates strong convergence between the quantitative and qualitative strands. Both highlight that socio-cultural constraints, provider behavior, and inadequate outreach combine to restrict youth service utilizations.

## Discussion

The findings indicate that youth access to sexual and reproductive health (SRH) services is constrained by structural, socio-economic, and cultural determinants. Globally, accessibility challenges are linked to poverty, mobility constraints, and facility distribution (10). WHO emphasizes proximity, affordability, and flexible service hours as core standards for youth-friendly services (11). In this study, limited service hours, centralized facilities, and absence of youth-specific spaces discouraged utilization, particularly among disadvantaged youth. Decentralizing SRH services through schools, community centers, and outreach programs has been shown to increase utilization (12, 13), supporting a multi-level approach combining health system reform, community engagement, and policy resource allocation.

Confidentiality concerns were widely reported. Quantitative findings showed perceived lack of privacy, while qualitative data revealed structural limitations, provider breaches of trust, and community stigma. Addressing confidentiality requires structural reforms such as private counseling spaces and soundproof rooms (14), alongside behavioral change through continuous provider training. Rights-based health systems depend on providers internalizing confidentiality as both a legal and moral obligation (15). Trust-building must therefore be institutionalized within youth-centered service delivery.

Provider attitudes significantly affect service use. Unfriendly and judgmental behaviors discourage adolescents from seeking care and reduce repeat visits (14,16). Sustained capacity-building, supportive supervision, mentoring, and accountability mechanisms are essential to reinforce youth-friendly standards (17). Integrating youth feedback systems such as satisfaction surveys and community scorecards further strengthens accountability and mutual respect (14).

Financial barriers also limit access. Low-income communities face compounded exclusion due to poverty, distance, and weak infrastructure (18). Transport and opportunity costs can represent substantial burdens. Evidence supports subsidized outreach services, school-based delivery, and voucher or cash-transfer schemes to reduce these costs (19). Intersectoral collaboration between health, education, and social protection sectors is therefore critical (20).

Youth-centered service delivery must move beyond rhetoric to active participation. Youth corners and participatory feedback platforms increase satisfaction and retention (14, 17). When youth co-design services, trust and agency improve (21). Strengthening school–health facility linkages and peer education models enhances knowledge dissemination and normalizes SRH discussions (19). Absence of mobile outreach and community dialogue reduces engagement (22), while stigma and provider moralism continue to discourage care-seeking (14,18).

Overall, improving youth SRH requires a multi-sectoral strategy addressing structural inequities, provider behavior, financial accessibility, youth empowerment, and community norms to ensure equitable and sustained utilization of youth-friendly SRH services The regression analysis shows that age significantly predicts perception (F = 3.967, p = 0.047), with a small but statistically significant negative effect (B = –0.189, β = –0.098, p = 0.045), indicating that perception slightly decreases as age increases **(Table 5).**

**Table 5.** Relationship between age and SRH perception (simple linear regression result).

| Model |  | Unstandardized Coefficients |  | Standardized Coefficients | t | Sig. |
| --- | --- | --- | --- | --- | --- | --- |
|  |  | B | Std. Error | Beta |  |  |
|  | (Constant) | 2.738 | 0.113 |  | 24.166 | 0.000 |
|  | Q1.1. What is your age in years | -0.189 | 0.094 | -0.098 | -2.013 | 0.045 |

Although the model explains a meaningful portion of the variance, a substantial residual (SSE = 188.598) suggests that other factors also influence perception; overall, age has a statistically significant but modest impact on the outcome **(Table 6).**

**Table 6.** Relationship between father education and SRH perception (Linear regression result)

| Model |  | Sum of Squares | df | Mean Square | F | Sig. |
| --- | --- | --- | --- | --- | --- | --- |
|  | Regression | 1.847 | 1 | 1.847 | 3.967 | .047b |
|  | Residual | 188.598 | 405 | 0.466 |  |  |
|  | Total | 190.446 | 406 |  |  |  |

## CONCLUSIONS

The study’s findings indicate youth not starting from zero, there is clear awareness that SRH information and shared responsibility matter and many youths express a strong desire to learn more. yet the overall knowledge environment remains fragmented, superficial and heavily shaped by schools, media and peers rather than a balanced mix that includes families and trusted health professionals.

At the service-delivery level, YFSRH is not experienced as reliably youth-friendly, structural barriers including distance, inconvenient service hours, lack of dedicated youth spaces and costs combine with quality barriers such as insufficient privacy, uncertainty about confidentiality, limited educational materials, weak feedback and accountability mechanisms and perceived judgmental or culturally insensitive provider interactions to reduce trust, discourage repeat visits and reinforce the pattern of reactive care-seeking.

Most importantly, qualitative insights suggest that even when providers understand and promote rights-based, participatory care, these principles are not consistently operationalized in day-to-day service environments, leaving a gap between policy intentions and lived experience.

## Authors’ Contributions

GG, SK and DM were responsible for conceptualization, investigation, methodology, writing, review and editing and funding acquisition. GG was responsible for data collection. for the formal analysis. SK and DM were responsible for supervision

## Authors’ Information

## Data Availability

All data set are available upon request

## Acknowledgments

I sincerely thank the Guraghe Zone Health Department, Guraghe Zone Education Department, Yaberus Preparatory School, District Health Offices, health facilities, and all study participants for their cooperation, facilitation, and invaluable contributions to this research. I also gratefully acknowledge the University of South Africa (UNISA) for providing bursary support for this study.

